# Design of logistics Indicators for Monitoring the Covid-19 Vaccination Process in Colombia

**DOI:** 10.1101/2023.02.23.23286347

**Authors:** Nubia Velasco, Andrea Herrera, Johanna Trujillo-Diaz, Ciro-Alberto Amaya, Catalina González-Uribe, Estefania Hernandez

## Abstract

The vaccine supply chain (VSC) integrates all the activities from production to dispensing. It is characterized by its complexity and low responsiveness, highlighting the importance of defining key performance indicators (KPIs). The design science research methodology was used to develop 38 KPIs, which were grouped into humanitarian and technological dimensions. The former includes demographic, epidemiological, and vaccination indicators, while the latter is classified into five groups according to the echelons of the supply chain. Public and private health organizations and research groups validated the indicators. They were calculated and recorded daily to monitor the logistics progress of the National Vaccination Plan against covid-19 in Colombia. These tailored KPIs, made it possible to evaluate and compare the results of the execution and effectiveness of public policies, and to redefine the strategies, showing that the logistics point of view helps identify the impact of good practices and transfer them promptly.

## 1. Introduction

The vaccine supply chain (VSC), like other supply chains, integrates all the necessary activities and interactions from the production process until the vaccines reach the population. According to de Carvalho et al. [1], the VSC involves five levels, from suppliers to vaccinated individuals. The first two steps are included in the manufacturing process, while the last three are part of the delivery process. Once a vaccine is produced, it is shipped for international distribution and stored under specific conditions. The last step is the delivery to different customers (countries) where, according to specific policies, it is distributed to the final consumer. This research aims to establish KPIs to evaluate the performance of VSCs in the particular case of covid-19 vaccination in the context of the National Vaccination Plan (PNV by its acronym in Spanish) in Colombia to provide suggestions to support decision-making and improve vaccination operations.

Before the covid-19 pandemic, VSCs were already considered complex due to three main factors: product complexity, globalization, and regulation [1], and thus, a proper reconciliation of economic, technological, and value key performance indicators (KPIs) is required to identify the real impact of vaccination programs [2]. Regarding their last mile, Duijzer et al. [3] demonstrated that the increasing number of studies on VSCs reflects the importance of logistics in the success of vaccination campaigns. Additionally, Shah [4] identified the need to improve the responsiveness of VSCs. Specifically, the author states that in the case of an emergency such as bioterrorism or fast-developing epidemics, the responsiveness of VSCs is slow, taking up to 24 months from vaccine development to reaching consumers [5]. Furthermore, Zaffran et al. [6] identified the necessity of strengthening the VSC in different dimensions as information management, human resources, and optimization, among others; nevertheless, most of the recent works have been developed for the optimization of supply chain [7] and Lemmens et al. [8] in their literature review on supply chains models, identified key issues specifically for VSCs, one of them being performance measures. Subsequently, Lemmens et al. [2] proposed a general framework for defining KPIs for VSCs, considering that these supply chains should support access to medicines. To our knowledge, this is the first and only framework designed for VSCs, and the authors proposed three dimensions of KPIs: economic, technological, and value or humanitarian. Firstly, the economic dimension includes measures such as cost, profit, economic value added, and net present value, widely known concepts determined by internal factors and by external aspects such as competition, government, and donor policies. Secondly, the technological dimension represents the performance of the supply chain in terms of resource utilization, capacity, lead time, throughput, and vulnerability to uncertainties in the supply, such as demand (coverage), and in operations, such as responsiveness, volume, and delivery flexibility, and reliability, among others. Finally, the value or humanitarian dimension refers to the ethical, social, and individual values related to VSCs. Examples of indicators in this dimension are vaccine availability, number of fully immunized people, equity, and disability-adjusted life years (DALY).

Furthermore, at the end of 2019, covid-19 emerged, and in March 2020, a coronavirus disease pandemic was declared. Immediately, along with measures to prevent its spread, vaccination appears as a strategy to counteract it. The process of vaccination against covid-19 (CVP) has several objectives: a) to reduce the risk of death and prevent a severe course of the disease; b) to maintain essential health services and their infrastructure available; c) to protect employment and the economy [9]. To achieve these objectives, countries developed mass vaccination plans that can be viewed as a supply chain directly involving entities in the upstream and downstream flow of products and information [10]. When CVPs started, the supply strategy was limited to allocation focused on reducing mortality and morbidity rates, with priority given to age-group vaccination [11, 12]. Supply was the bottleneck [12, 13], as vaccines were in low supply and high demand [14], making the application of the first doses slow [11]. Supply, distribution, and logistics in the CVPs were the ones that marked the flow and speed of vaccination [15], and Colombia was no exception. The covid-19 VSC in Colombia, proposed by the Ministry of Health and Social Protection (MSPS by its acronym in Spanish), was based on the Expanded Immunization Program (PAI by its acronym in Spanish). To increase capacity and minimize the risk of waste [16], the PNV supply chain has a five tiers structure [17]: (1) global vaccine manufacturing laboratories, (2) a central warehouse (located in Bogotá), (3) departmental or municipal health secretariats, (4) health service provider institutions (IPSs by its acronym in Spanish), and (5) vaccination centers. Figure 1 represents the Colombian covid-19 VSC. On the left, the macro-logistics is represented with Bogotá in the center of the Colombian map, the second tier, and the 37 territorial entities representing the health secretariats. On the right side, the micro-logistic process is depicted.

**Figure 1.**
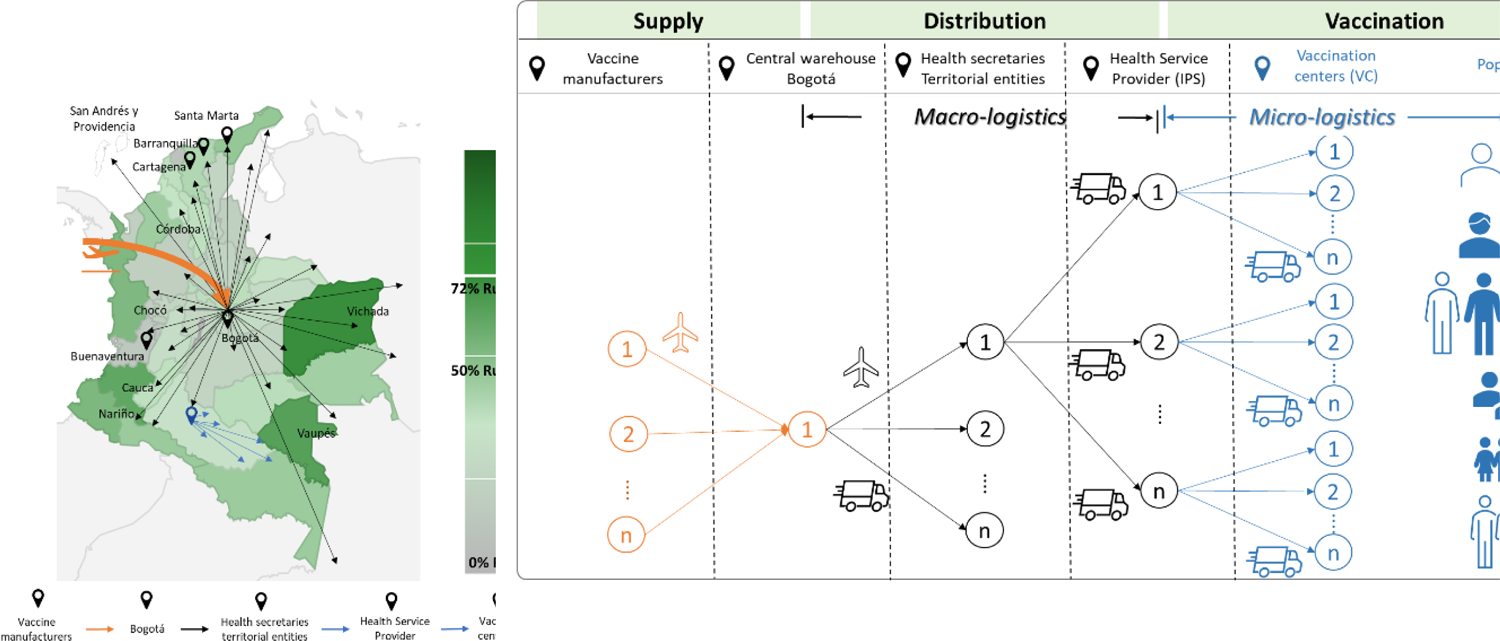
Colombian covid-19 vaccine supply chain[17].

The allocation and distribution of vaccines to territorial entities were made according to an effectiveness criterion that depends on the vaccine availability, the doses allocated, and the vaccines administered [18]. However, these indicators do not reflect the specificities and peculiarities of the territorial entities, and they do not suggest the impact of logistics activities on the performance of the PNV.

To identify VSCs indicators and, specifically, those used in the covid-19 VSC, we conducted a review of covid-19 vaccination dashboards that we classified according to three objectives: a) to track reactions in patients vaccinated against covid-19 [19]; b) to monitor vaccination progress by a geographical location (worldwide, regional, or local) [9, 19–22], and c) to track operations in vaccination centers [19]. In the second and third groups, more than 30 key metrics were found to measure geographic coverage. These KPIs were grouped by similarity and are summarized in Table 1.

**Table 1.**
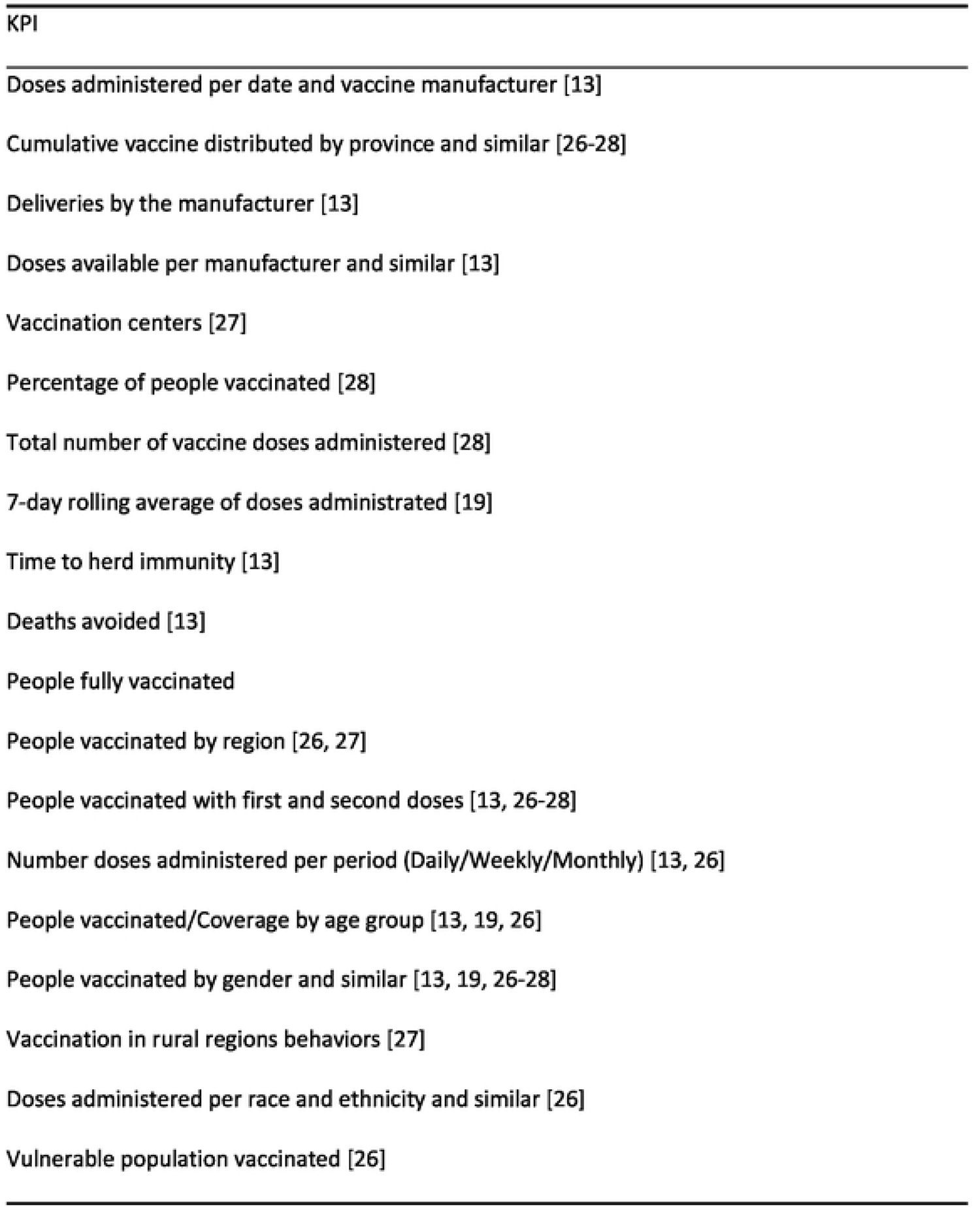
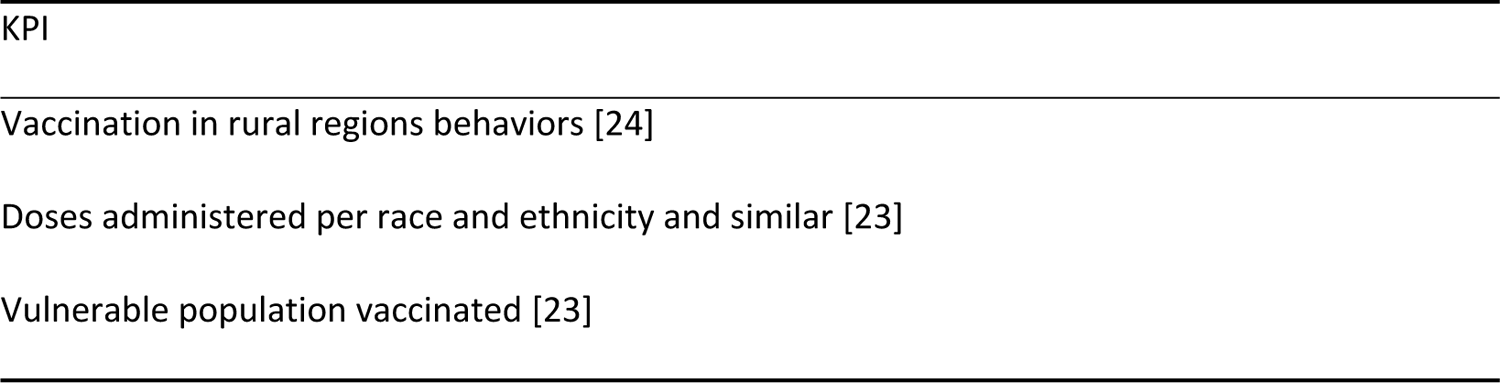
Indicators published in dashboards and articles.

As shown in Table 1, most KPIs are defined for the humanitarian dimension, but few are related to the technological dimension of VSCs. To the best of our knowledge, there is no study aimed at charting the daily progress of the covid-19 vaccination process in terms of the VSC.

## 2. Materials and Methods

We used the design science research (DSR) approach to identify KPIs and design the dashboard, which has proven helpful in this context [13, 17]. DSR is a problem-solving paradigm that seeks to build a working artifact (i.e., the dashboard). Its relevance is based on the applicability of the artifact in practice (i.e., monitoring the vaccination process to enable faster response). DSR is an iterative improvement methodology, and each iteration is composed of six stages [17] (1) problem identification and motivation, (2) goal definition and solution, (3) design and development, (4) demonstration, (5) evaluation, and, (6) communication.

The KPIs proposed are based on available structured and unstructured official information sources We included all available structured and unstructured official information sources to design the KPIs, as shown in Table 2. Unstructured official sources include the MSPS Twitter, vaccine allocation resolutions, and MSPS infographics, while structured ones include demographic data by territorial entities, open data on vaccination progress and vaccine arrivals, and open data on SARS-CoV infection.

**Table 2.**
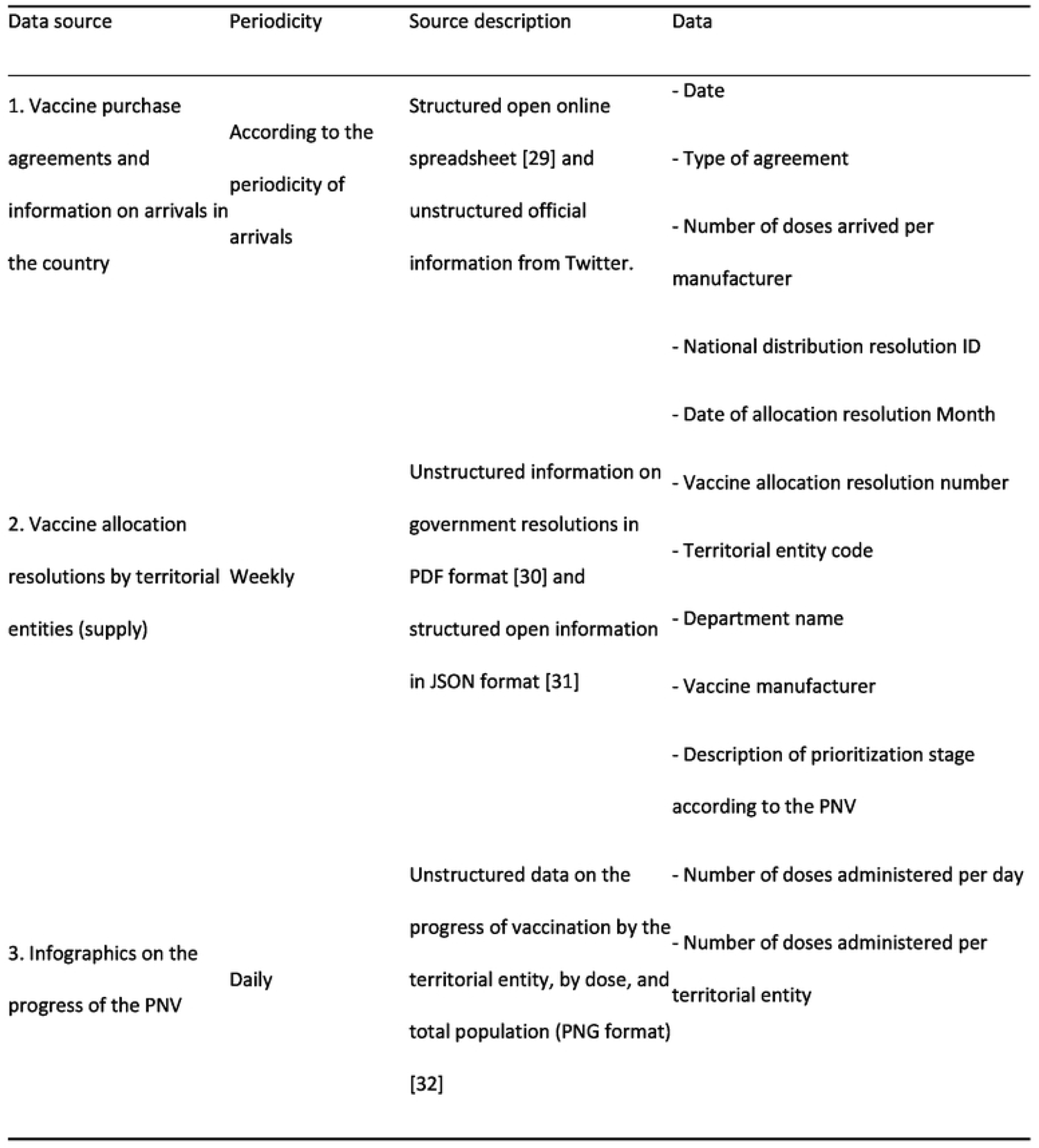
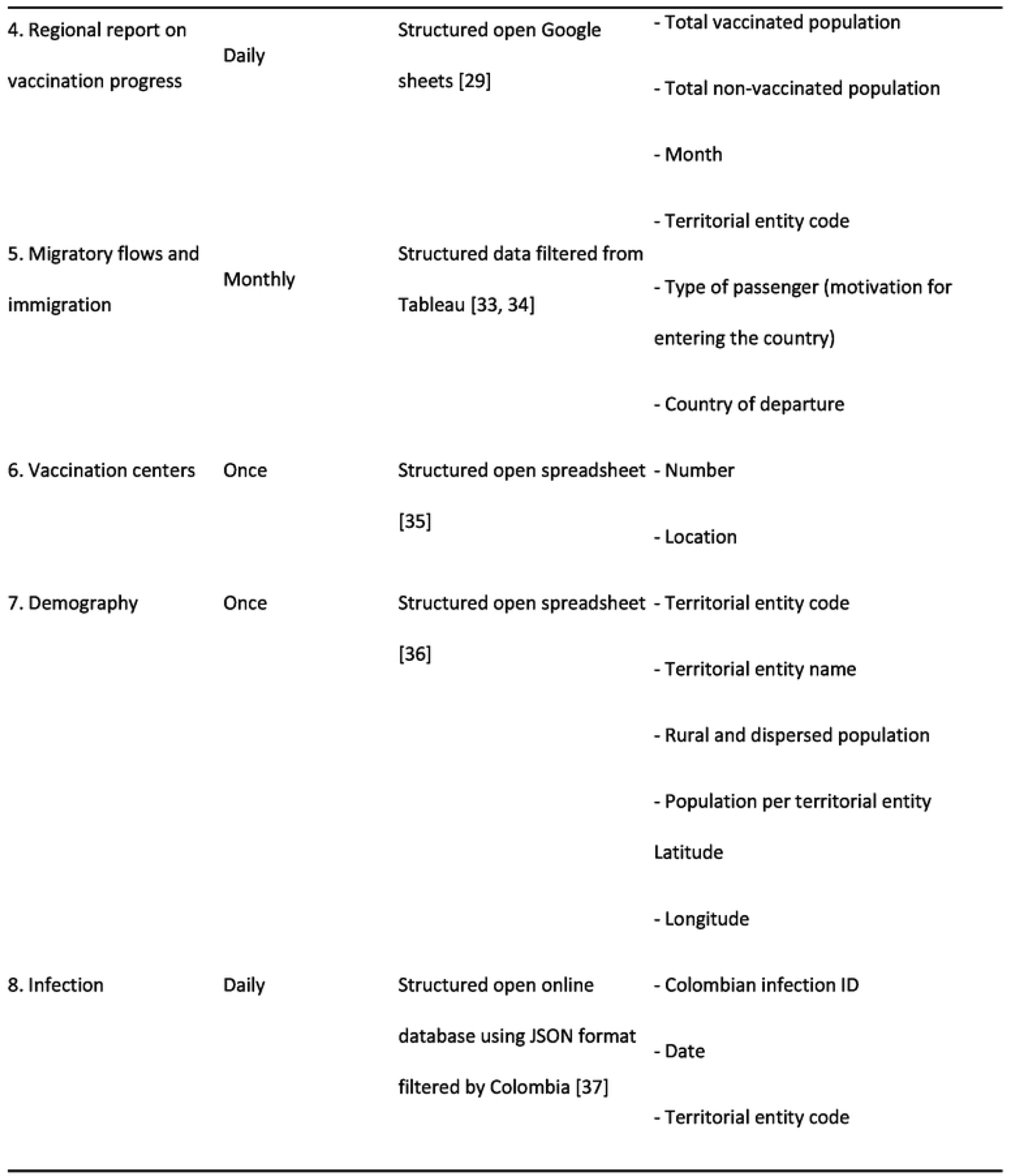
Structured and unstructured data sources

Based on the above data sources and after validation with stakeholders of the health ecosystem in Colombia, 38 indicators were defined [35]. Some of them were adapted from the literature, and others were proposed to consider the characteristics of the covid-19 vaccination process: massive, with different types of vaccines and involving two doses and boosters, among others. Economic metrics were not considered due to a lack of information about vaccination costs and because, in the case of covid-19 vaccination, the purpose was to vaccinate people quickly. Following the classification proposed by Lemmens et al. [2], we grouped the KPIs into the humanitarian and technological dimensions. In the humanitarian dimension, in addition to the KPIs related to vaccination (C), we included epidemiological indicators for covid-19 (A) and demographic indicators (B) to identify the territorial entities with the greatest need and the level of efficiency of their processes. The technological ones are classified into five groups, according to the echelons in the supply chain: (D) supply and distribution, (E) capacity, (F) coverage, (G) forecasting, and (H) logistics. The latter was included, considering the need to track vaccine availability and establish allocation and distribution policies. Table 3 shows the indicators for each group, and Appendix A contains the equation that defines each indicator.

**Table 3.**
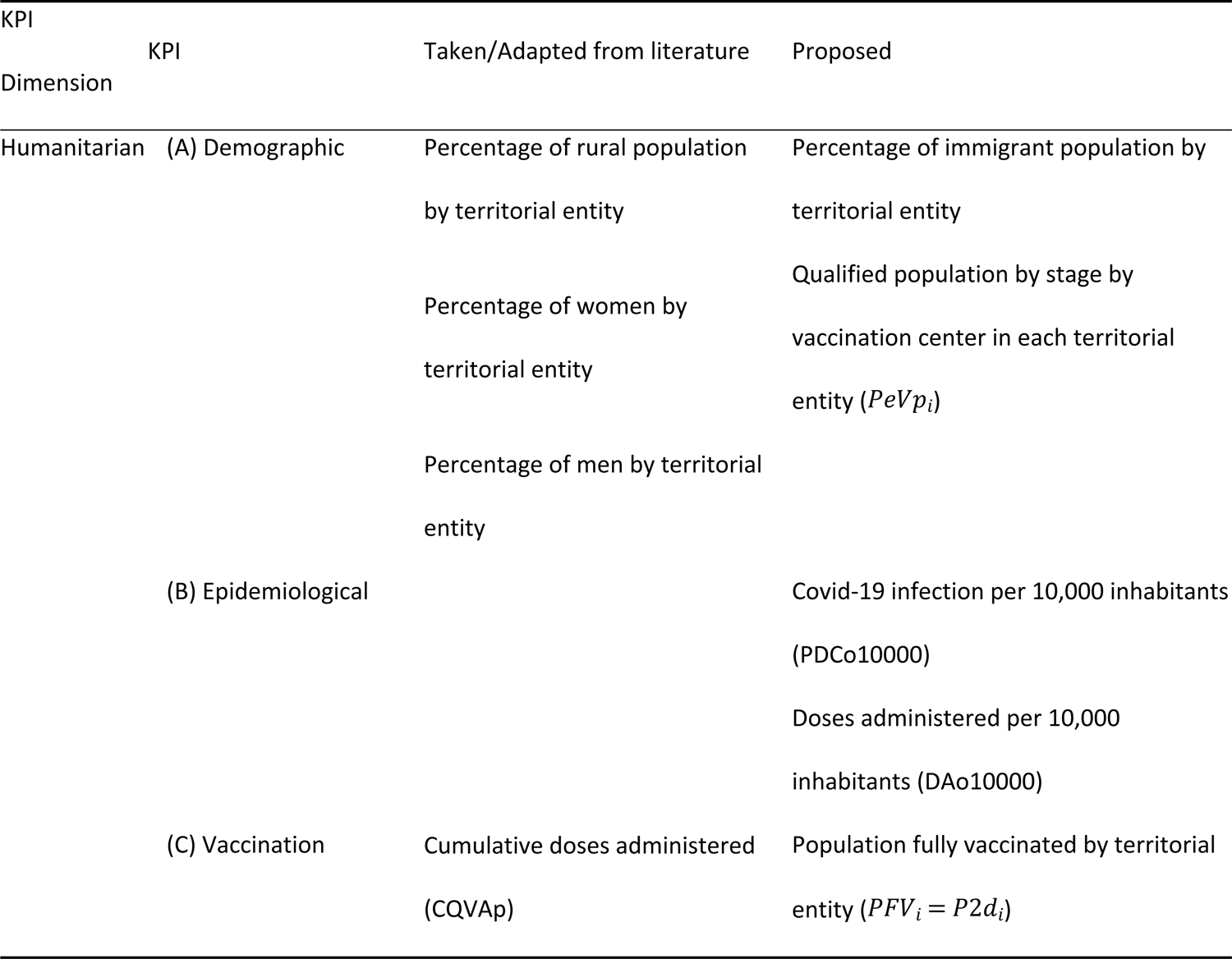
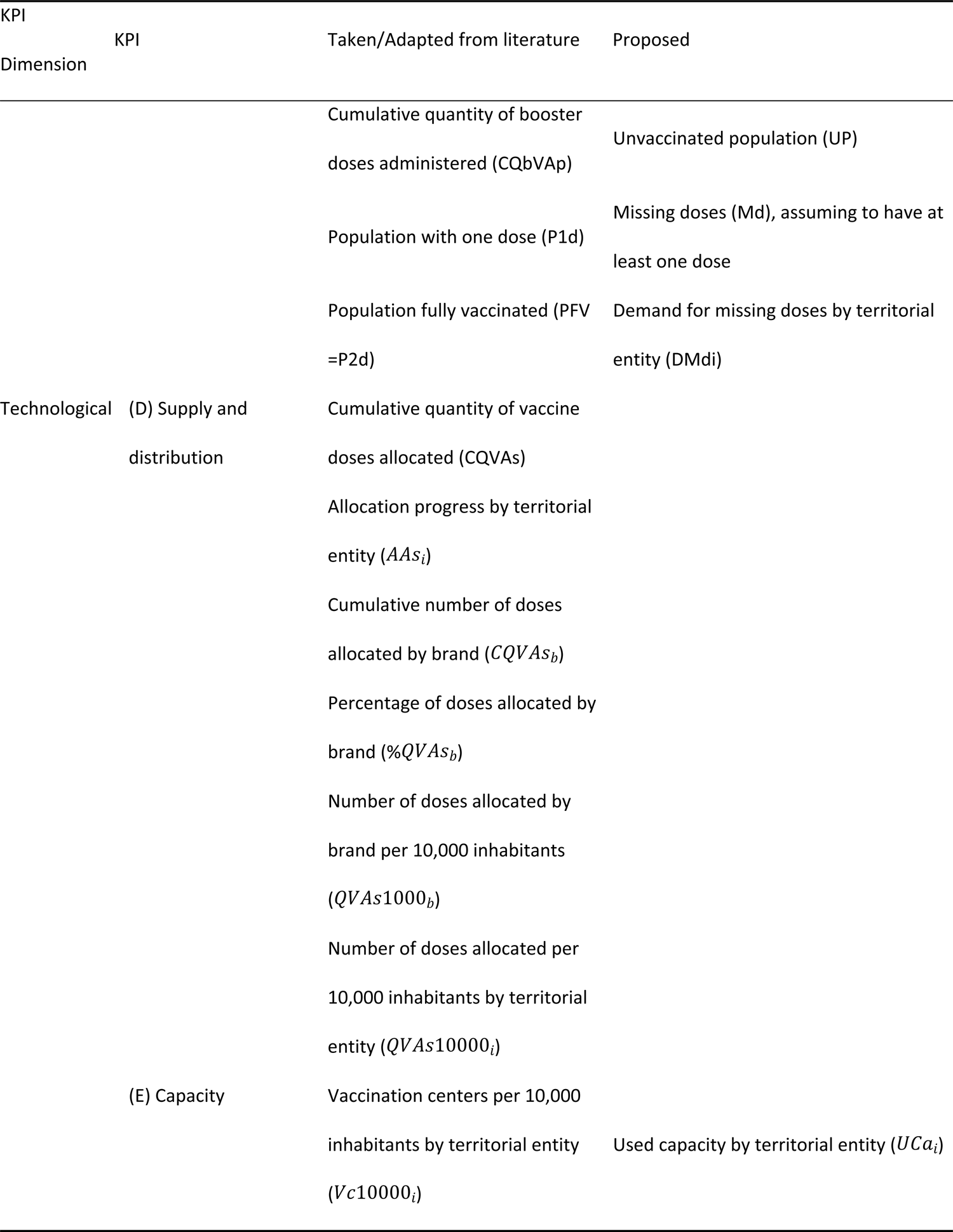
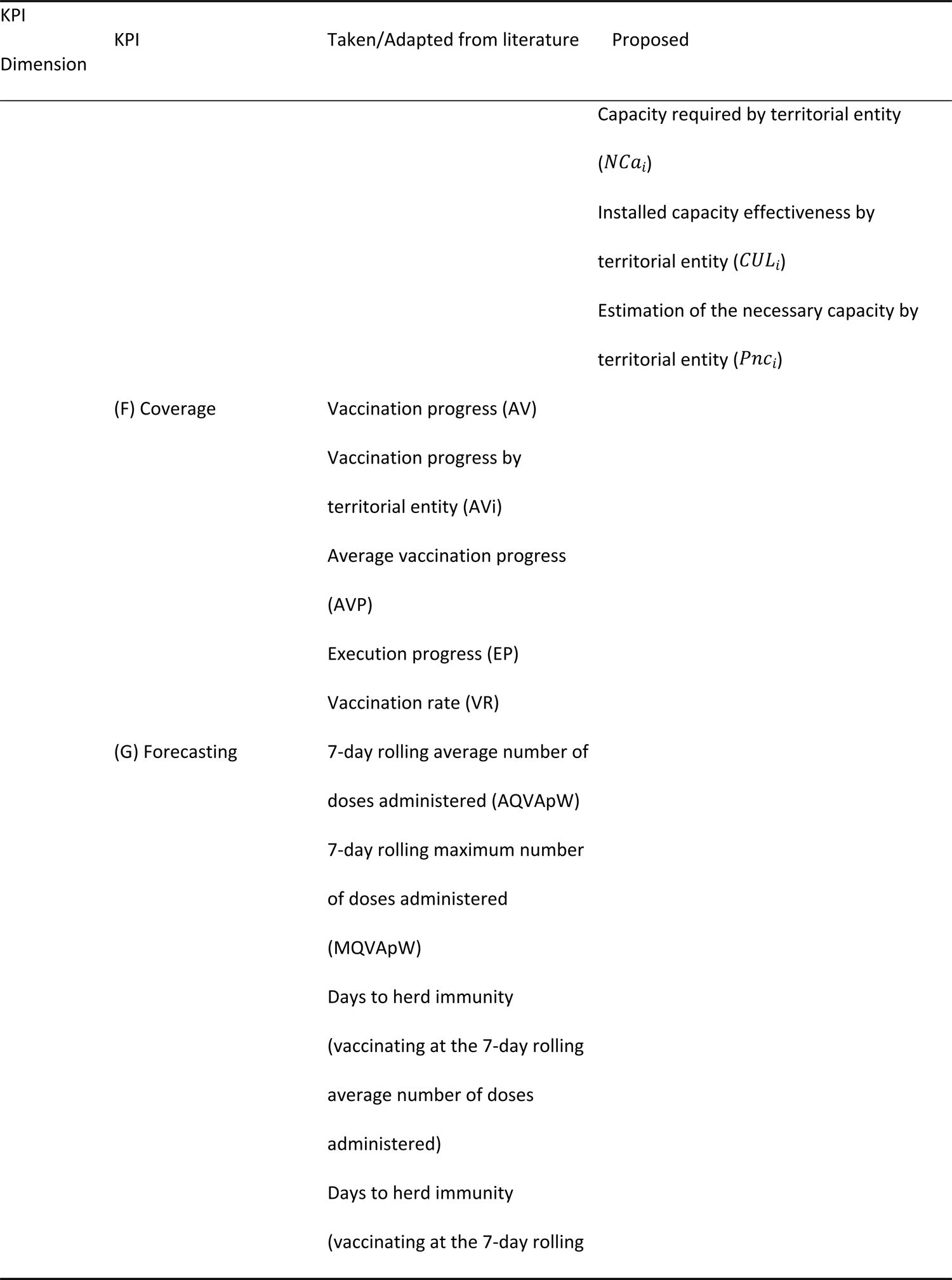
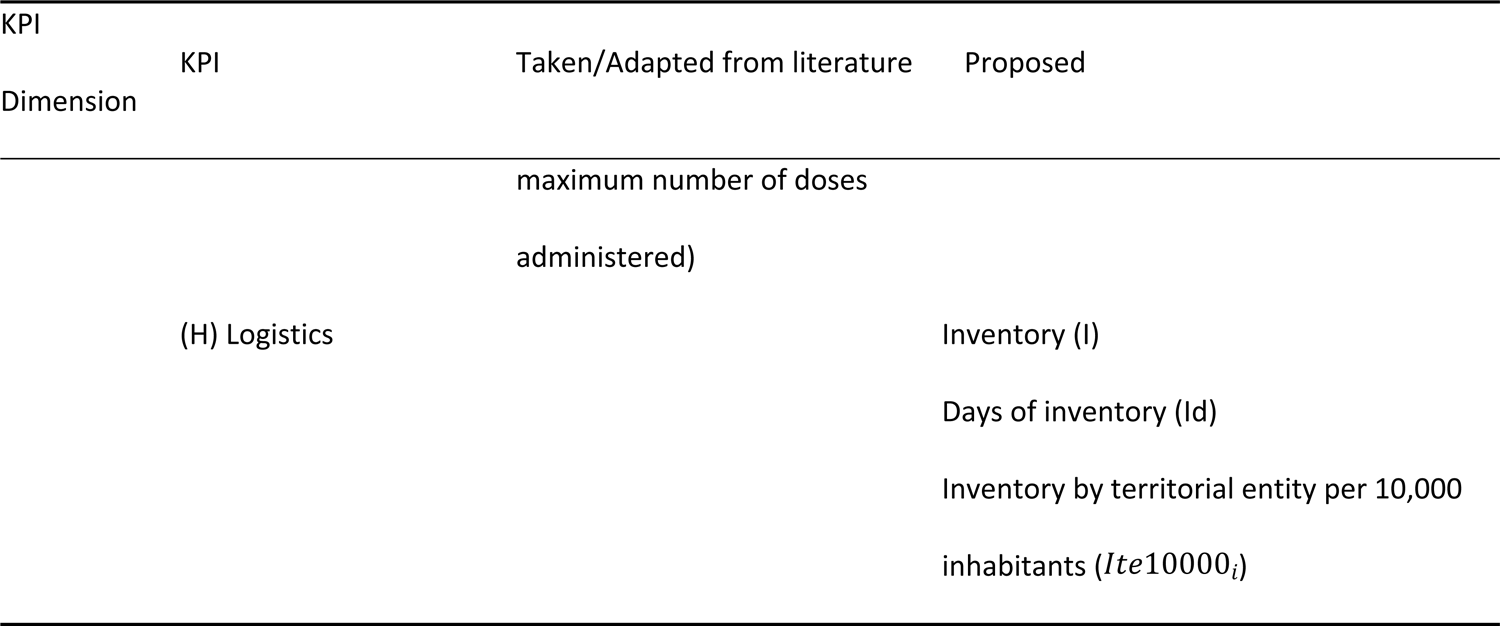
Indicators designed and validated through iterations.

A snowflake data model was defined to manage the data sources and generate all the indicators (see Appendix B). The snowflake is composed of a fact table (called time series) that includes the eight groups of indicators and seven dimensions tables: (1) international vaccines purchase agreements and arrivals, (2) demographic information, (3) national distribution, (4) vaccination progress, (5) covid-19 infection, (6) migration information, and (7) time; and their attributes [36]. Data were daily collected and the KPIs were generated by department, major cities, and by stage of the PNV.

Since the objective is to track the logistics progress of the PNV, we developed a Google Studio dashboard divided into three parts: (a) PNV progress and projections, (b) PNV logistics KPIs, and (c) capacity and efficiency of vaccination centers; progress was daily tracked starting on 17th of February; with the defined goal being to vaccinate at least 70% of population (herd immunity) by December 31st, 2021. To better communicate the progress of the PNV to stakeholders, we generated a monthly newsletter summarizing the progress and the main leading suggestions derived from the analysis of the logistics KPIs. The newsletter was composed of seven sections: a) population to be covered; b) progress by geographical entity; c) vaccination rate; d) comparison among doses assigned, administered, and inventory; e) supply and demand analysis; f) estimated completion date; g) available inventory by population [37]. The indicators and the dashboard were socialized with the departmental and municipal health secretaries to support their decision-making associated with the PNV. The first dashboard was published on April 22nd, 2021. The last newsletter summarizing the results of the PNV was shared with the MSPS and other stakeholders in March 2022 [38].

## 3. Results

The 38 proposed KPIs were used to track the logistics progress of the Colombian PNV from February 16th to December 31st 2021, the time window defined by the MSPS to vaccinate 70% of the population (i.e., administering more than 70M doses). Each day, information published in different sources was manually uploaded to a database (see Table 2), which automatically computed the KPIs according to the snowflake data model (see Appendix B). The calculations were displayed in a dashboard developed on Google Studio, available on the COLEV website (https://colev.uniandes.edu.co/22-colev-en-accion/192-avance-nacional-de-vacunacion-2). The main results by KPI group are discussed below.

Regarding demographic indicators, the PNV was modified throughout 2021 to include more population. Thus, by December 31^st^ 2021, 95% of Colombian inhabitants were eligible to be vaccinated, including those over three years of age and regular and non-regular migrants. 36% of the population belongs to stage five (between 11 and 39 years of age), 14,4% are between 3-11 years of age, followed by stage 1 with 12,6% of the population. The higher concentration of people is located in Bogotá, Antioquia, Valle del Cauca, and Cundinamarca, the country’s center.

According to the National Administrative Department of Statistics (DANE by its acronym in Spanish) [33], departments such as Vichada and Vaupés have the highest percentage of rural population with 72% and 65%, respectively, followed by Cauca (62%), Nariño (55%), and Choco (54%). In these departments, a different strategy may be needed to achieve the PNV goal and cover the population.

In addition, epidemiological indicators show that at the end of 2021, covid-19 infections per 10,000 inhabitants (Figure 2(a)) were mainly reported in the center of the country (darker red), apart from the departments of Amazonas and Atlántico, which are considered departments with high entry/exit of people and trade. This KPI reflects where the covid-19 transmission was more intensive, indicating regions where the vaccination needed to be accelerated.

**Figure 2.**
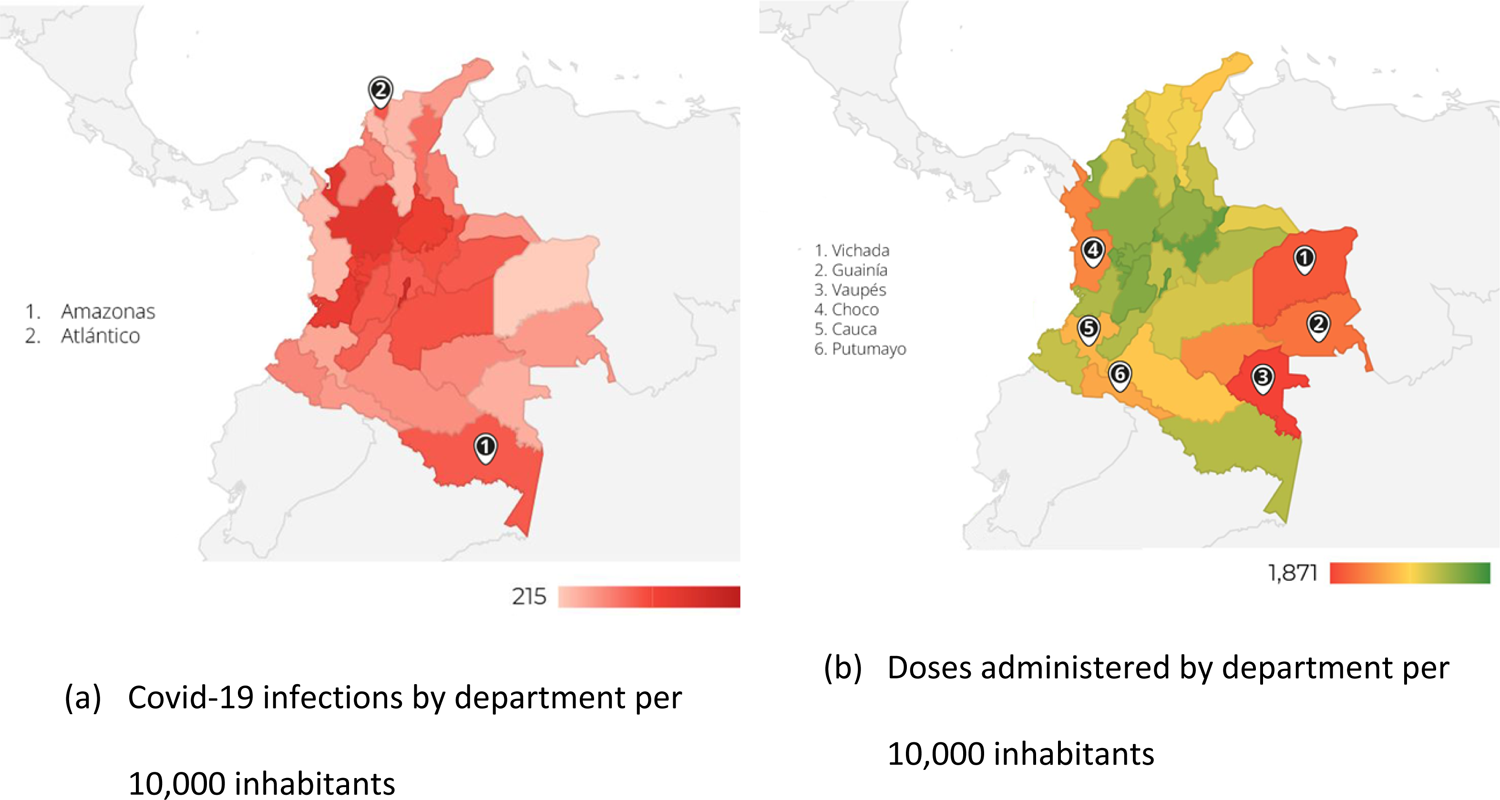
Epidemiological KPIs.

Figure 2(b) shows the doses administered per 10,000 inhabitants up to December 31st. Departments in the center of the country have a higher allocation, while the eastern departments present the lowest allocation. Additionally, some of the departments in orange (Chocó, Cauca and Putumayo) in the western part of the country are departments with geographical areas with significant socioeconomic inequalities.

Regarding vaccination results, the PNV reached 55.5% (28.3M) of fully vaccinated population, 19.6% (10M) with one dose, and 6.5% with a booster dose (3.3M) [29], surpassing countries such as Russia, Poland, Bulgaria, Bolivia, Venezuela, and Nicaragua [34]. As of December 31st, there was an 18.4% of unvaccinated population.

Regarding the monthly vaccination progress, represented by the cumulative doses administered per month (Figure 3), there was an increase in the cumulative number of doses administered from May to August, explained by the opening stages and by the administration of second doses, which were the highest proportion from July to December (green bars). The population that started the vaccination scheme was constant from August to December (orange bars – Population with one dose). In July, Colombia began to dispense a mono-dose vaccination scheme, which also contributed to accelerating the immunization process, mainly from September onwards (green line representing the percentage of population fully vaccinated).

**Figure 3.**
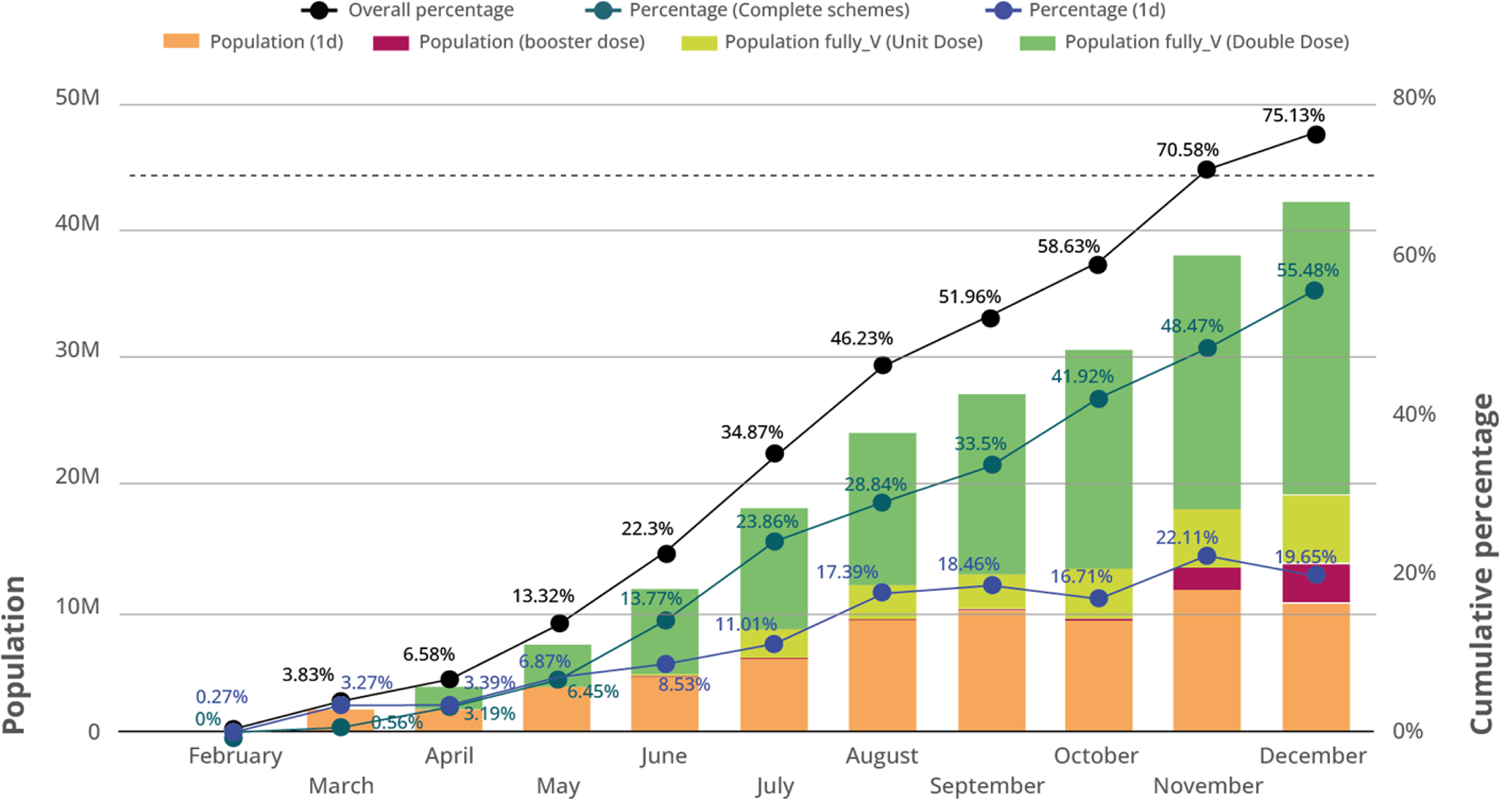
PNV monthly progress.

Figure 3 shows how people vaccinated with second doses (green bars) began to replace those who started the vaccination scheme (orange bars), representing half of all doses administered from September to December. It is important to note the increase from October to November due to the expansion of the qualified population to be vaccinated (children and migrants) and the requirement of a vaccination card to access public spaces.

Regarding supply and distribution, vaccines from different laboratories were administered in Colombia. From February to June, the vaccines allocated were mainly Pfizer and Sinovac. In March and April, some doses arrived from AstraZeneca. In June, Colombia received the first batch of Janssen, which made it possible to administer the mono-dose vaccine scheme. The country received Moderna vaccines through direct negotiation and donation from July to December. At the end of the year, Sinovac (29,3%) and Pfizer (26,1%) were the most allocated vaccines, followed by AstraZeneca (17,8%), Moderna (17,6%) and Janssen (9,2%), the former mainly allocated to rural territories. Figure 4 shows the number of doses allocated by brand.

**Figure 4.**
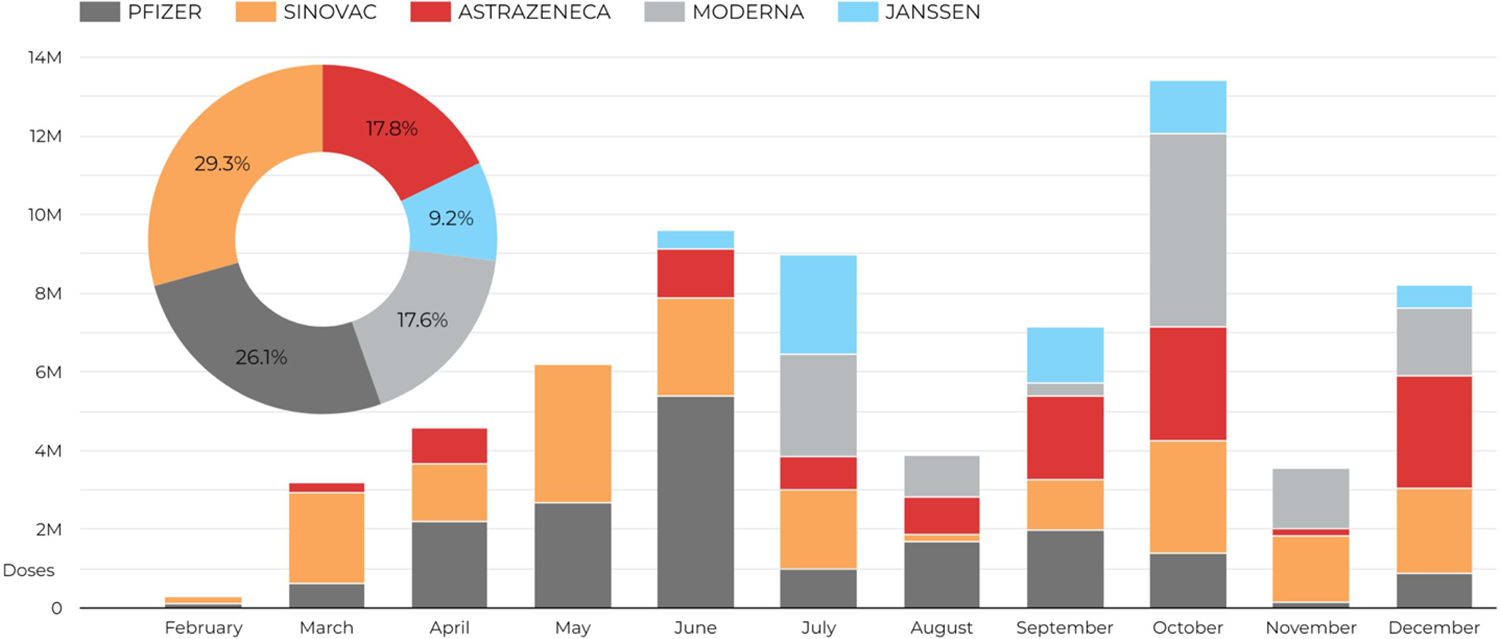
A cumulative number of doses allocated per brand.

Regarding the number of doses allocated per month, October was the month with the highest number of doses allocated, given the increased frequency of arrivals. In contrast, the same month presented a low vaccination rate (see Figure 4), reflecting the need to increase the population target.

In terms of capacity, Figure 5 shows the progress over the year of capacity required, and capacity used. The required capacity (red line) indicates the vaccination level per center needed to meet the PNV goal, while the used capacity (green line) indicates the number of doses administered per vaccination center. In general, the country was below the required capacity, except for few days during July and August, mainly due to the opening of stages 4 and 5. This result showed from the beginning that the country would not be able to administer the doses needed to achieve herd immunity.

**Figure 5.**
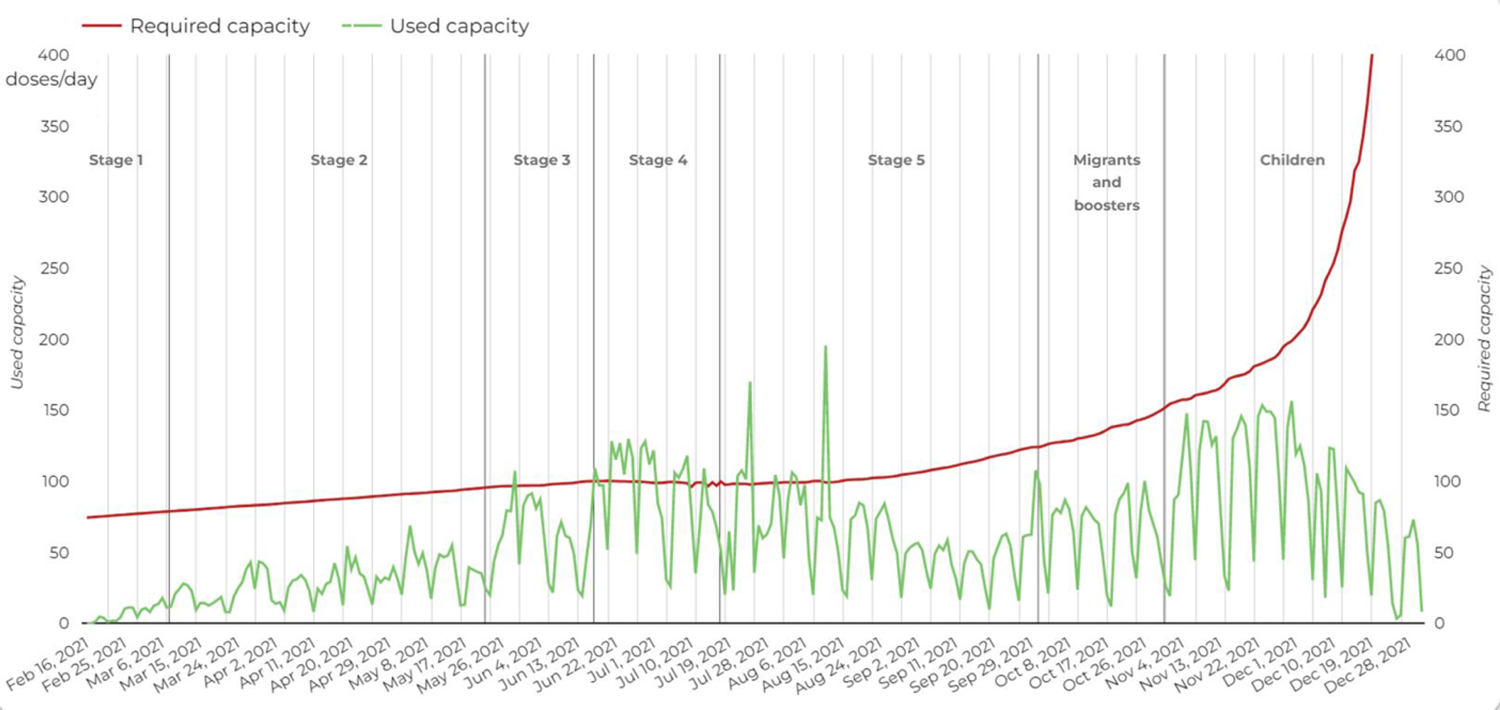
Analysis of the used and required capacity progress.

We made a comparative analysis among departments, using the qualified population (see Figure 6(a)), the number of vaccination centers per 10,000 inhabitants (see Figure 6(b)), and the installed capacity effectiveness (see Figure 6(c)), estimated as the ration between the used capacity and the required capacity (see Appendix A). Figure 6(a) shows that Vaupés is the department with the lowest population qualified to be vaccinated, with the highest number of vaccination points per 10,000 inhabitants and with the lowest installed capacity effectiveness (Figure 6(c)), then, Vaupés needs to better use its installed capacity. In contrast, Antioquia has the highest number of people qualified to be vaccinated, a small ratio of vaccination centers per 10,000 inhabitants, which are efficient used (Figure 6(b)), with an installed capacity effectiveness greater than one, indicating that their used capacity is greater than the capacity they need.

**Figure 6.**
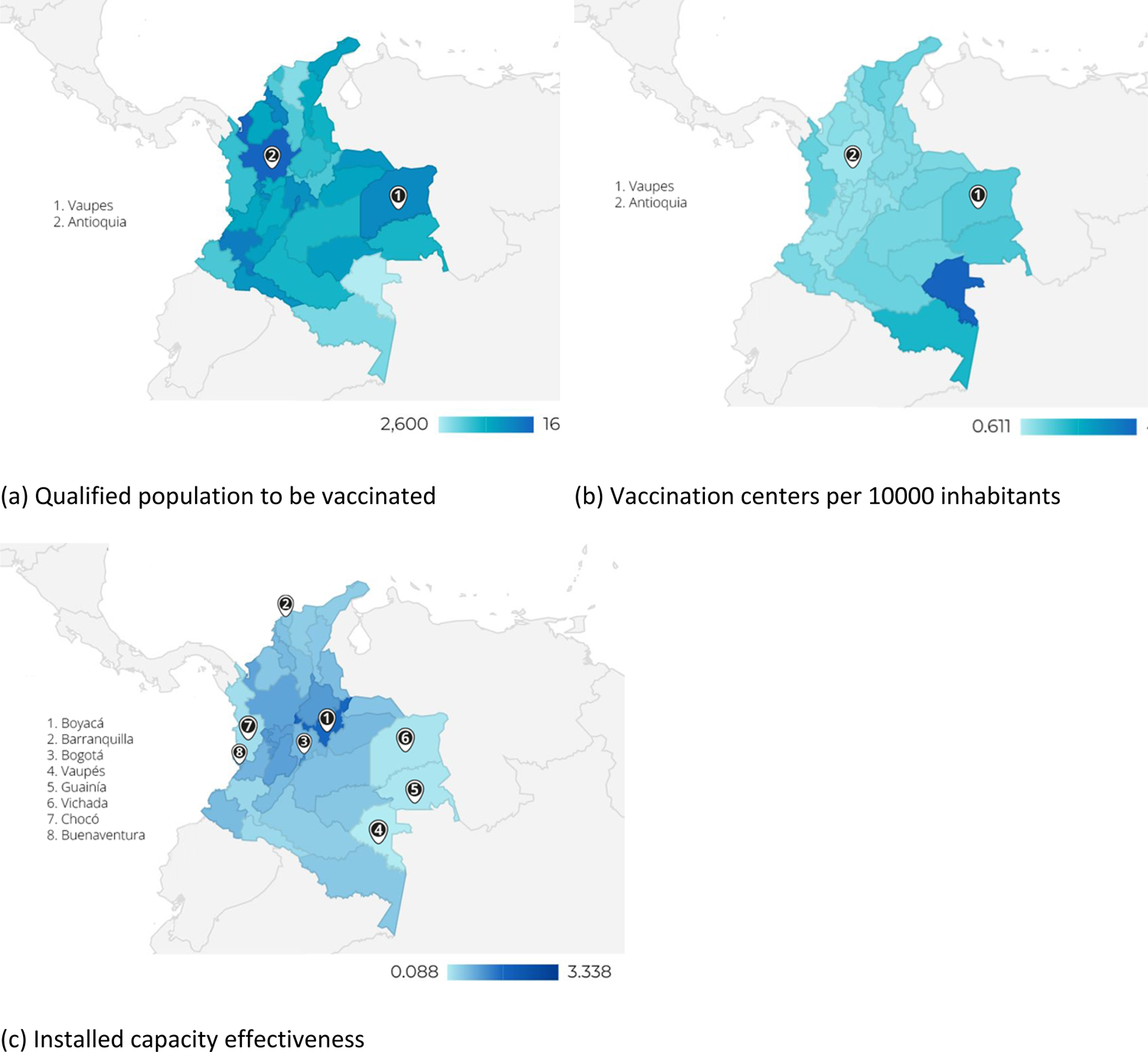
Capacity KPIs.

Considering the installed capacity effectiveness, the most efficient territories were San Andrés y Providencia (333%), Boyacá (170%), Barranquilla (167%), and Bogotá (115%) with capacity utilization of over 100%, indicating efficiency in the use of resources. While departments such as Vaupés (8,33%), Guainía (16,33%), Vichada (16,96%), Chocó (25,28%), and Buenaventura (29,74%) showed low efficiencies, mainly due to a low level of vaccine allocation (see Figure 2(b)).

With respect to coverage, after 318 days of PNV operation, 69M doses were allocated, of which 64.7M were administered [29]. At the beginning, between February 16th and May 21st, two stages were opened (out of five), at this time the daily vaccination rate was below the daily target (horizontal dotted line in Figure 7), set by the National Government at 250K doses. At the end of May, the government decided to accelerate the opening of the PNV stages, thus giving the opportunity to more population to access to vaccine. This action allowed increasing the daily vaccination up to 600K doses. However, for the fifth and last stage, the daily vaccination rate decreased rapidly, and by early August, this indicator fell below the target. Given this reduction, in October, vaccination was opened to the migrant population while the MSPS adopted booster doses. In addition, at the end of October, children aged 3 to 11 years were included in the PNV, and finally, at the end of November, to reactivate local businesses, a vaccine certification was required for access to public spaces. Thanks to these measures, the country reached 90% of the goal.

**Figure 7.**
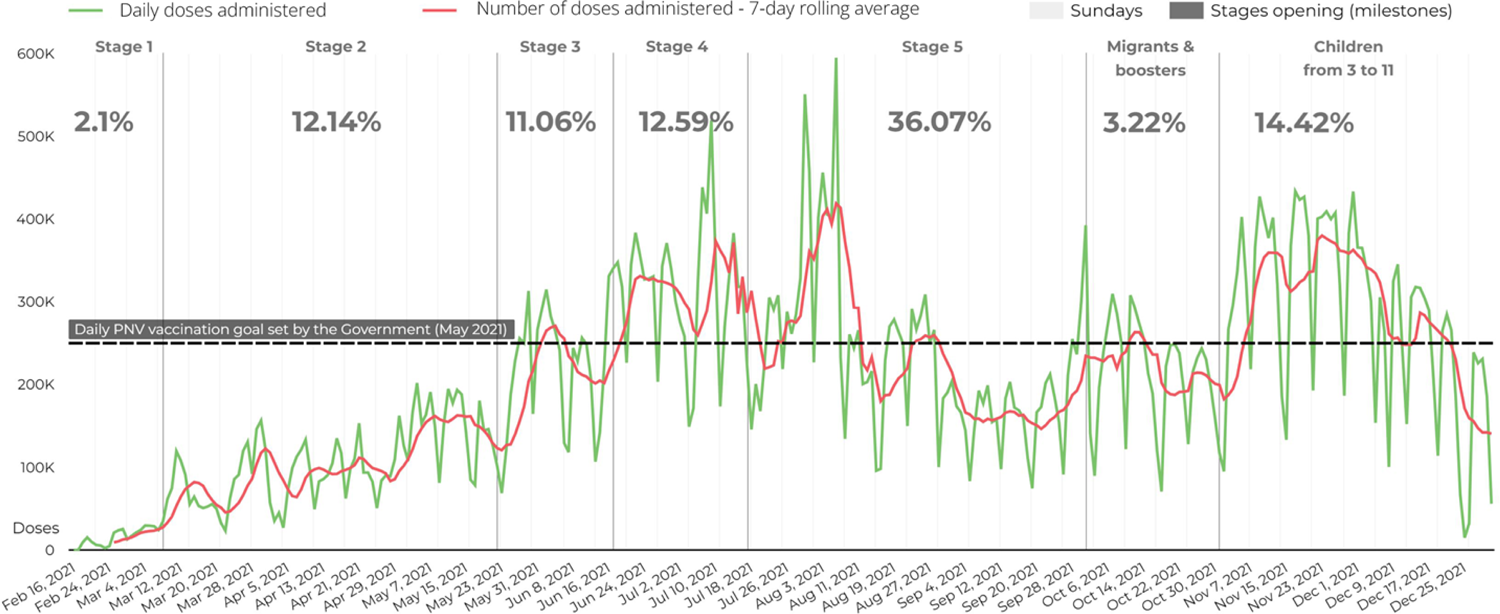
Daily vaccination rate.

Figure 7 shows that, throughout the vaccination period, Sundays were the day with the lowest vaccination rate. Additionally, the vaccination rate increased each time a new stage was opened, or a new government policy was implemented.

Concerning performance by territorial entity (Figure 8(a)), Barranquilla, Bogotá, and Cartagena, cities with large populations, exceeded 70% of the population vaccinated. Similar results were obtained by departments such as Risaralda, Caldas, and Quindío, small departments in terms of surface area. Although their population is rural, short distances facilitate distribution and coverage. Boyacá presented the best results, and the main reason is good management of vaccination progress. San Andrés y Providencia [39] and Amazonas [40] also exceeded 70% of the vaccinated population because a prioritization policy was applied given their geographic characteristics.

**Figure 8.**
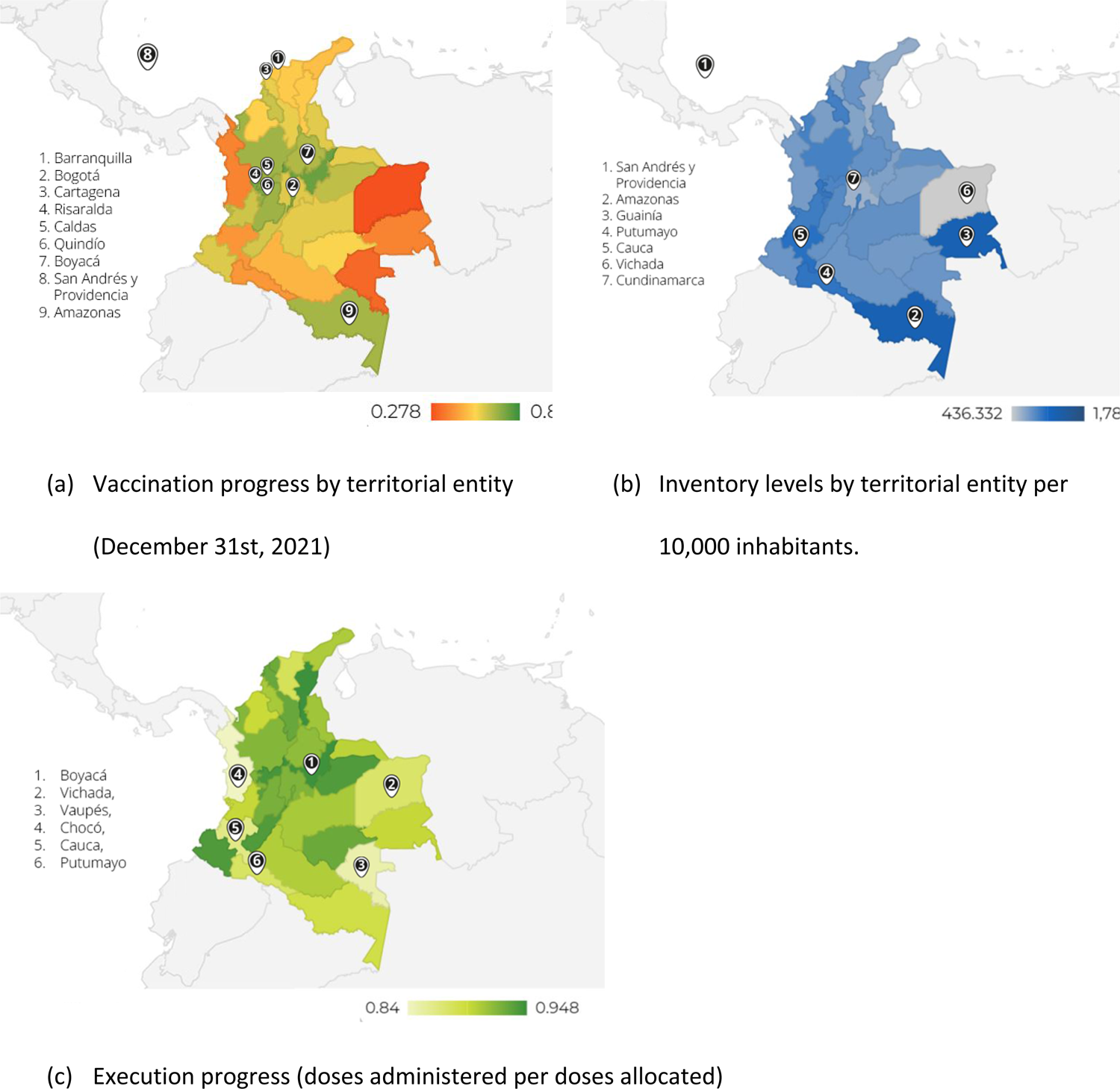
Vaccination progress, Inventory levels, and Execution

Regarding inventories, at the national level, 2021 ended with more than 12M vaccines in stock. San Andrés y Providencia (1,787), Amazonas (1,247), Guainía (1,245), Putumayo (1,012), and Cauca (1,040) were departments with the highest inventories per 10,000 habitants (see Figure 8(b)). At the same time, Vichada and Cundinamarca were the departments with the lowest inventory level, the former due to access difficulties and the latter due to efficiency in administering the allocated doses.

To estimate the remaining days to achieve the established target and thus herd immunity, the 7-day rolling average number of doses administered and the 7-day rolling maximum number of doses administered were used. In both cases and using information from the last week of December (i.e., 24th – 31st), Colombia would have reached 70% of the vaccinated population by January 21st, 2022. As of December 31st, territorial entities such as Antioquia, Barranquilla, Boyacá, Caldas, Cartagena, Bogotá, Quindío, Risaralda, San Andrés y Providencia, and Tolima had already met the goal, while Arauca, Atlántico, Buenaventura, Caquetá, Cauca, Cesar, Chocó, Córdoba, Cundinamarca, Guainía, Guaviare, La Guajira, Magdalena, Meta, Putumayo, Vaupés and Vichada would have taken more than three months to reach the goal, indicating the need to define new vaccination strategies for these territories.

Figure 9 shows the vaccine inventory (blue line), doses administered (green line), and doses allocated (green bars) every week since May 2021. The inventory level was 3M, and by the end of October, the inventory level reached 12M vaccines. Overall, Figure 9 shows that there was no shortage throughout the year; on the contrary, in October, inventory exceeded the allocation and administered vaccines.

**Figure 9.**
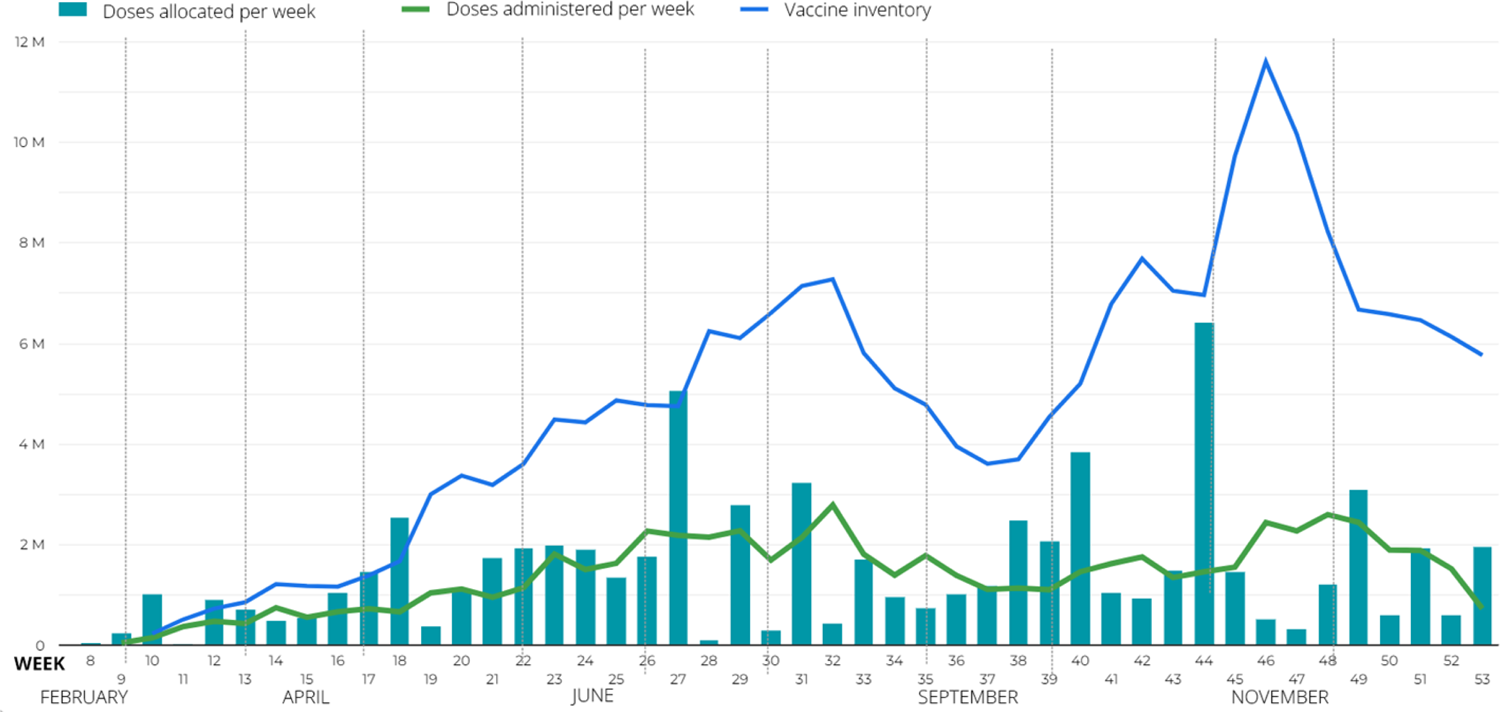
Inventory levels behavior.

We also conducted a combined analysis of indicators. To begin with, the progress (a coverage KPI – Figure 8(c)) shows that the most critical finding is that throughout the country, execution levels are high, the lowest being 84%, understanding the level of execution as the ratio between the vaccines applied over the vaccines assigned. Combining this result with the epidemiological KPIs (Figure 2), it is possible to monitor the speed of vaccination (doses administered per 10,000 inhabitants) versus infections and logistic execution (Figure 8(c)). For example, on December 31st, Boyacá presented the highest execution, a high number of doses administered to its population, and the number of infections was low, indicating that the strategy used to vaccinate was good. Vaupés, on the other hand, presented a low execution, with a low number of doses administered, but with few cases; in this department, as mentioned above, it may be necessary to define a strategy other than mass vaccination. The results presented are discussed in the following section.

## 4. Discussion

The proposed technological KPIs complemented the humanitarian KPIs, allowing for more in-depth monitoring of the VSC. The set of 38 indicators made it possible to evaluate the effectiveness of the Colombian Government in the decision-making process and to make recommendations on vaccination strategies differentiated by territorial entity. Although the progress of vaccination against covid-19 in Latin America and the Caribbean (LAC) has been uneven concerning reaching the national goal of vaccinating at least 70% of the population [41], Colombia demonstrated that its health system could respond to massive vaccination processes, thanks to rapid and dynamic epidemiological and logistic decisions. As illustrated, the country reached 75% of the vaccinated population with at least one dose, showing a good performance compared with other LAC countries [34].

In particular, the proposed indicators made it possible to trace the assertive epidemiological decisions made by the Colombian government to achieve the PNV goal, for instance: Study the behavior of specific strategies, such as the blocking vaccination strategy at borders [40] and some other territorial entities [42, 43]. This strategy was used in March 2021 in Vaupés, Guainía, Amazonas, and San Andres y Providencia to prevent the entry of the gamma variant to Colombia. The strategy prioritized the allocation of vaccines to these departments and unified the stages of the PNV, giving access to all qualified persons to be vaccinated. We validated the allocation, distribution, and inventory, with the proposed KPIs confirming that at least Amazonas and San Andres y Providencia were among the first regions to meet the goal.

Identify and eliminate access barriers. In February, the PNV established that each health insurance company was responsible for vaccinating its affiliated members. To this end, each company had to contact its members and inform them of their vaccination appointment (day, time, and vaccination center). This procedure constituted a barrier to access, and it was quickly eliminated (June, stage 3), effectively increasing the vaccination rate, as shown in Figure 7.

Expand the population eligible for vaccination. In May 2021, the MSPS decided to include the entire population over 16 years of age [44] as part of the fifth stage. Then, in June, children over 12 years of age were added to the same stage [45], and finally, in October, regular and irregular migrants [18, 46] and children over 3 years of age [47] were added. This policy had a positive impact on vaccination progress, as shown in Figure 5 and Figure 7. During June and July, the used capacity exceeded the required capacity, indicating that the PNV goal could be reached before December 31st. Although the used capacity was lower than the required capacity in October, these indicators still improved.

Imposing the vaccination card requirement in public spaces (restaurants, movie theaters, and shopping malls) in November. This policy was effective and helped increase vaccination rates during November, mainly in large cities such as Bogotá and Barranquilla and departments such as Antioquia. Nevertheless, were identified some fake cards.

On the other hand, logistic decisions such as signing contracts with multiple vaccine suppliers [11], seeking approval of different vaccines by the National Institute for Drug and Food Surveillance (INVIMA by its name in Spanish) [48], seeking vaccine donations, continuously promoting the opening of the vaccination centers [11, 49], and maintaining continuous communication strategies; had a positive effect on the VSC, increasing the capacities of the system avoiding a massive vaccine shortage.

The technological KPIs, particularly the logistic ones, allowed us to identify that the VSC designed for Colombia had high inventory levels, reaching 12M vaccines by October 2121, which meant that at the end of the year, demand had to be boosted to reach the goal established by the PNV (see Figure 9). Additionally, at the end of 2021, vaccination progress of 23,8% was identified in rural areas such as Vichada, Vaupés, Chocó, Cauca, and Putumayo, which also had the lowest allocation progress. On the contrary, in large cities such as Barranquilla, Bogotá, and Cartagena, both allocation and vaccination improvement were high, reaching more than 70%. This situation, among others, is due to the lack of indicators to adequately allocate vaccines and make decisions using other variables such as the vulnerable population or the number of vaccination centers per 10,000 inhabitants (inventory, capacity, or coverage KPIs).

Concerning the allocation KPIs, from Figure 2 is possible to identify that the departments located in the center of the country have a higher allocation. In contrast, the eastern departments presented the lowest allocation. This could be explained by the fact that the center of Colombia is the most populated region, while departments such as Vichada, Guainía, and Vaupés (red on the map) are not. Comparing the two maps in Figure 2, it is observed that the departments with the highest number of cases are the same ones with the highest number of doses administered, which is good because it is crucial to be efficient in regions with an increased number of infections. However, departments such as Vaupés and Vichada, with a low concentration of cases and the highest percentage of rural population, have the lowest level of doses administered, indicating the importance of defining different strategies to reach the people in these regions. Comparing the allocation progress (figure 2(b)), the administration progress (figure 8(a)), and the execution percentage (Figure 8(c)), it is possible to identify that Vichada, Vaupés, Chocó, Cauca, and Putumayo have the slowest progress of allocation with a high level of execution (up to 80%). At the same time, these departments have inventory levels close to 20% of vaccines (see Figure 8(b)) and high levels of rural populations (see Figure 4), showing the efficiency of the vaccination process in rural territories.

According to some studies, the more vaccine supplied, the higher the willingness to be vaccinated [11], and herd immunity could be achieved quickly. In July, the fifth stage was opened, representing more than 30% of the qualified population. Consequently, the inventory levels decreased (see Figure 9). However, the vaccination rate also fell in August due to vaccine shortages. Following this situation, vaccination levels did not increase as expected, and mandatory vaccination policies had to be implemented to increase willingness to be vaccinated. Governments should ensure better provision in vulnerable areas and deploy extra-mural, home-based, or itinerant vaccination strategies to achieve better coverage.

Colombia’s PNV goal was to vaccinate 70% of the population by December 31st. Although this goal was not met, the designed VSC administered close to 70 million doses in less than a year, compared to the 5 million vaccines the health system routinely administered, representing a 14-fold increase. This result demonstrates that the Colombian health system can comply with a massive vaccination plan such as that of covid-19, supported by its logistics capacity and a flexible and dynamic policy.

The more information is open, available, accessible, and captured in real-time, the lower the uncertainty [13]. In the case of the mass vaccination process in Colombia, access to information could have been more timely and with different data sources and formats. As a consequence, some of the traditional KPIs of the VSC could not be included, such as a) the level of compliance with the negotiation (quantities purchased vs. quantities received, which is part of the supply), b) the wastage of covid-19 vaccines; c) the real-time capacity of the VSC; d) the real-time inventory per tier; e) the backlog; f) the doses administered in vulnerable population by age or ethnicity [19]

## 5. Conclusion

This research was developed in partnership with local public and private decision-makers, who validated the different phases of the project as well as the proposed KPIs and results obtained. This kind of collaboration helped consolidate a dashboard with different perspectives on the progress of the vaccination process in Colombia and to provide holistic feedback to health system stakeholders.

In this paper, we present 38 KPIs for the VSC. These KPIs were classified into eight groups: demographic, epidemiological, vaccination, supply, and distribution, capacity, coverage, forecasting, and logistics. The first three groups belong to the humanitarian dimension, and the others to the technological dimension. The proposal is based on a literature review identifying the need for technological KPIs for CSV before the covid-19 pandemic. KPIs were used to monitor the NVP in Colombia from February 16th to December 31st, the time window defined by the Colombian government to meet the goal of fully vaccinating 70% of the population. The results show that by combining technological KPIs with humanitarian KPIs, a comprehensive understanding of the whole process is achieved, making it possible to evaluate the effect of public health policies or even anticipate their behavior. During the development of this research, the authors identified the great need for access to open, standardized, updated, and official information.

## Author Contributions

N. Velasco: Conceptualization, Formal analysis, Methodology, Project Administration, Resources, Validation, Visualization, Writing—original draft preparation, Writing—review and editing, Supervision A. Herrera: Methodology, Formal analysis, Validation, Visualization, Writing—review, and editing, Supervision J. Trujillo: Data curation, Formal analysis, Software, Investigation, Validation, Visualization. C-A. Amaya: Formal analysis, Validation, Visualization, Writing—review, and editing, Supervision C. Gonzalez-Uribe: Funding acquisition, Formal analysis, Review and editing E. Hernandez: Data curation, Visualization All authors have read and agreed to the published version of the manuscript.

## Institutional Review Board Statement

The study was conducted in accordance with the Declaration of Helsinki and approved by the Ethics Committee of Universidad de los Andes (Acta: No.1394 – 2021).

## Data Availability Statement

The data will be available in the data repository of the Universidad de los Andes: https://papyrus-datos.co/dataverse/uniandes_grupo-de-investigacion_colev.

## Data Availability

All data files will be available in the data repository of the Universidad de los Andes: https://papyrus-datos.co/dataverse/uniandes_grupo-de-investigacion_colev.

## Acknowledgments

This research was developed in partnership with local decision-makers and health organizations. We thank the Ministry of Health and Social Protection for the interviews to present the validation results of the logistics monitoring board. We also thank Dr. Gabriel Carrasquilla G., Así Vamos en Salud (AVS), and the committee of health secretaries.

## Appendix A Indicators to trace the PNV

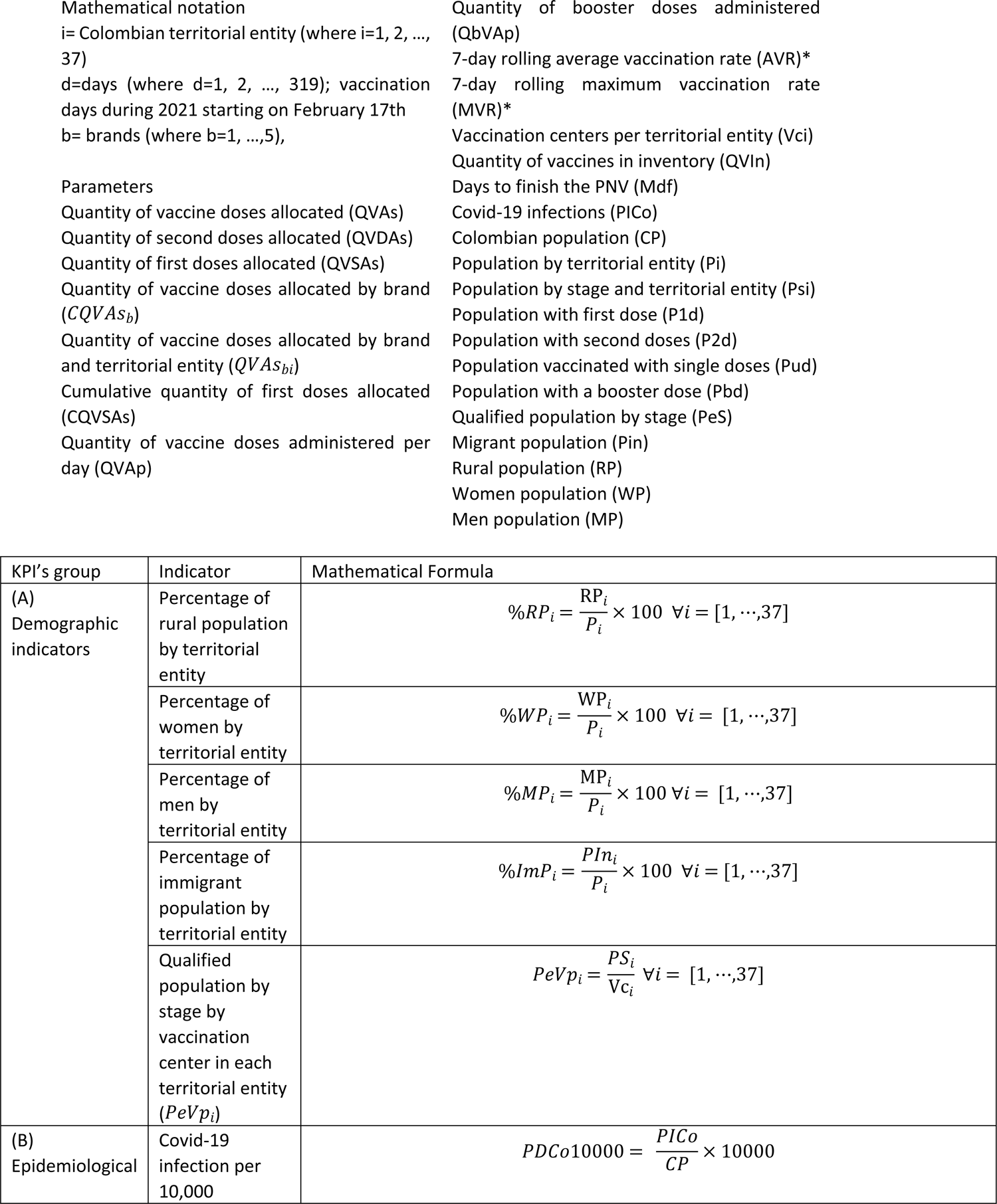
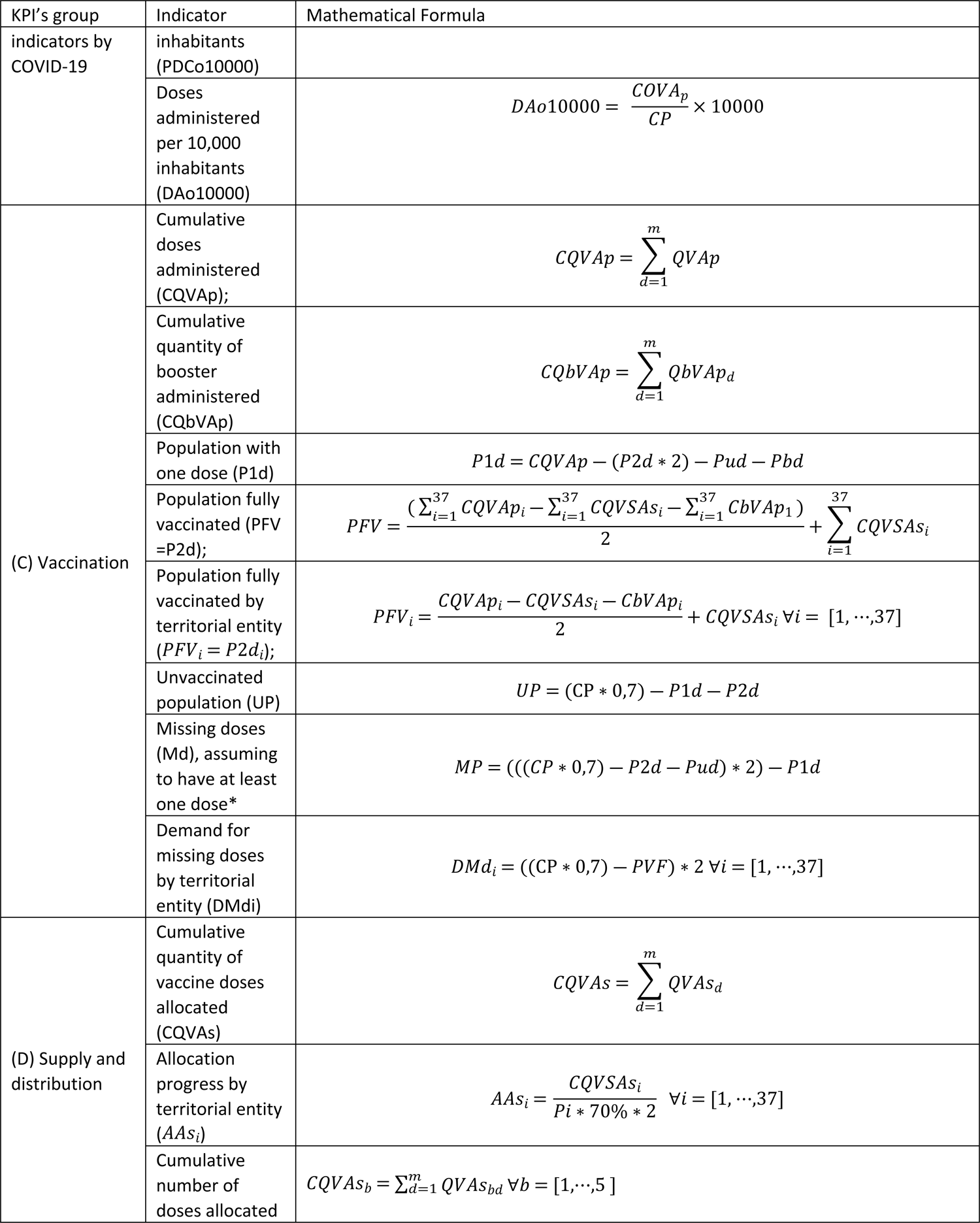
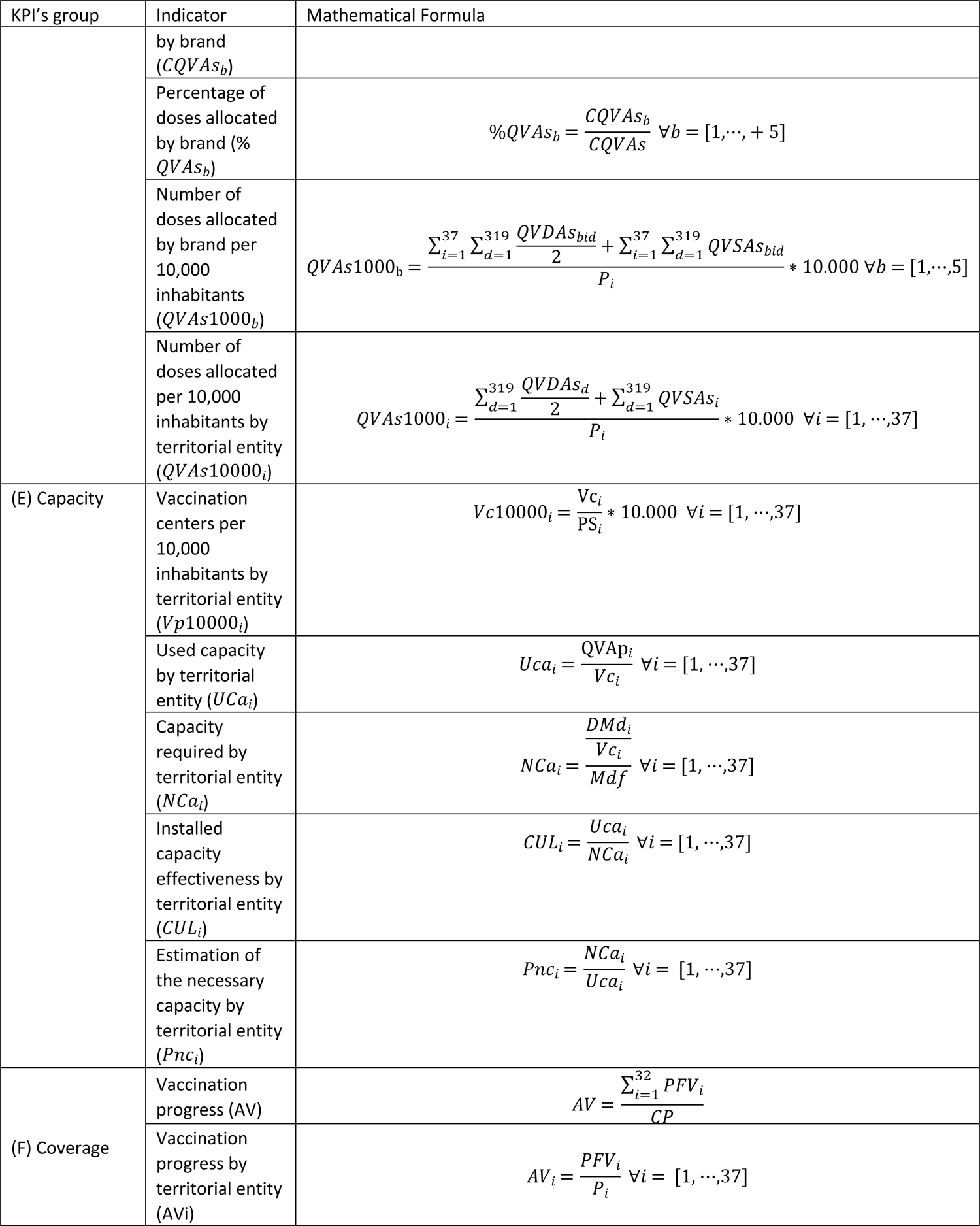
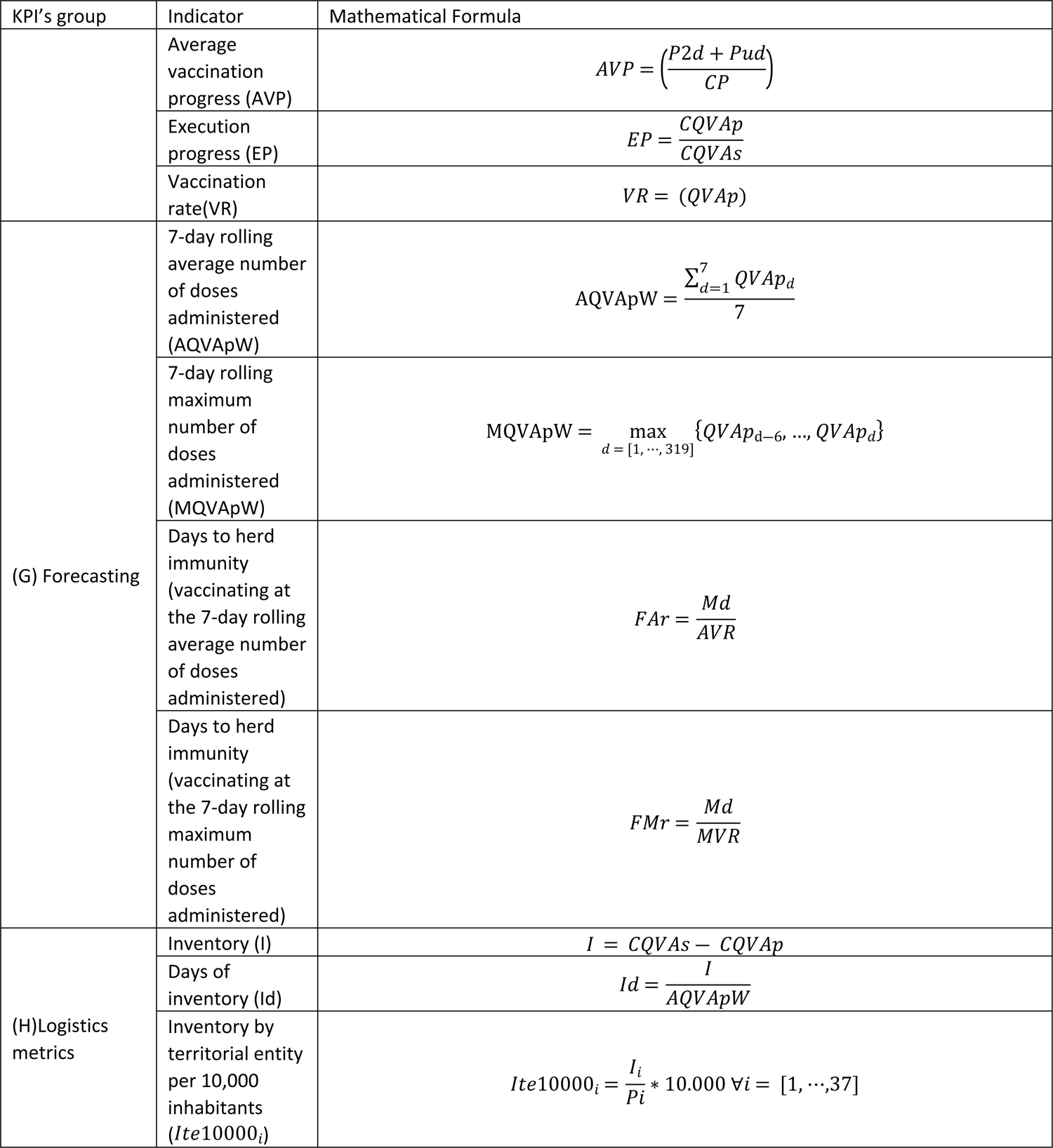

## Appendix B Snowflake dashboard data model

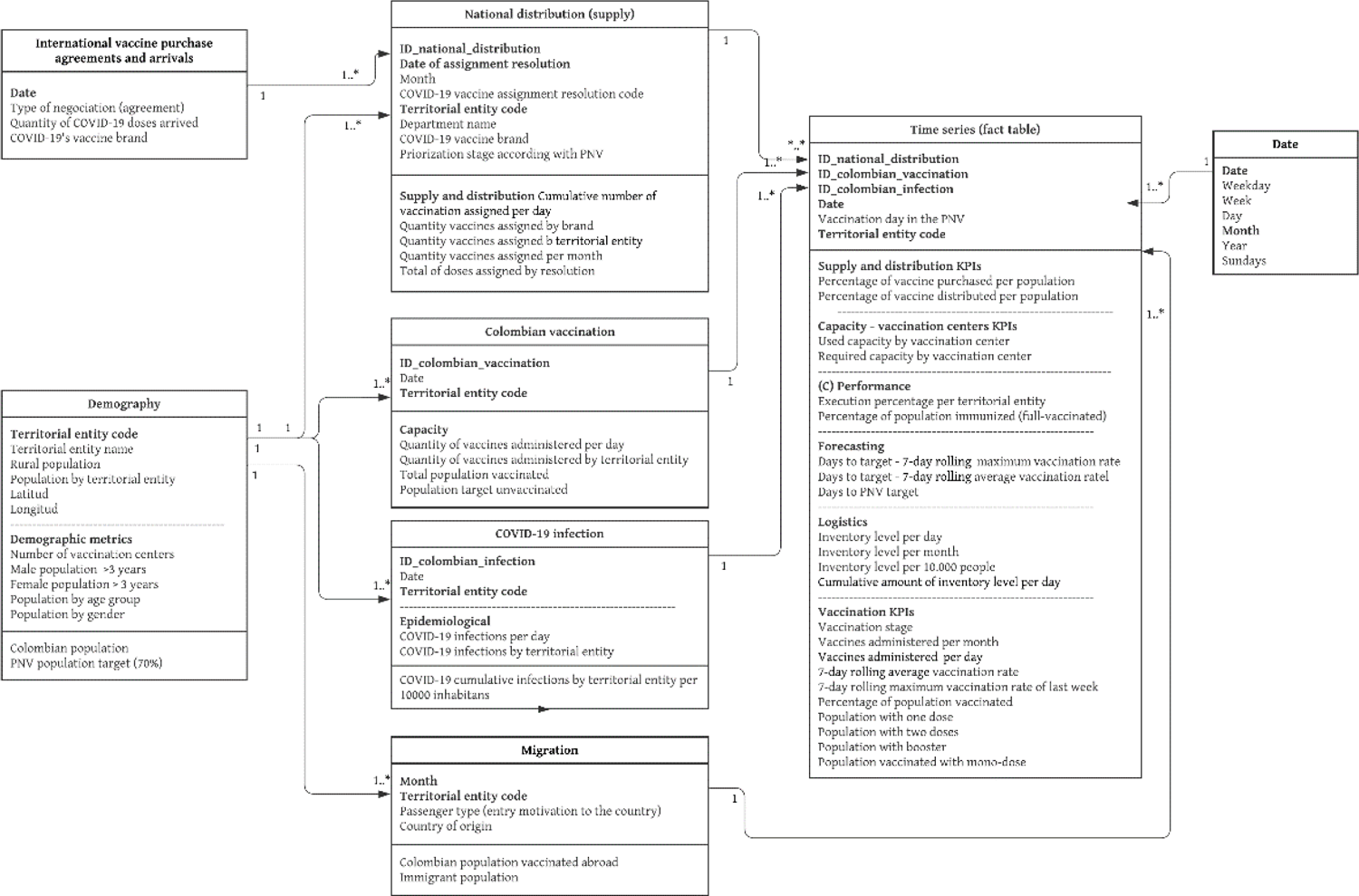

